## Supplementary Figure for "Genome-wide analyses of neonatal jaundice reveal a marked departure from adult bilirubin metabolism"

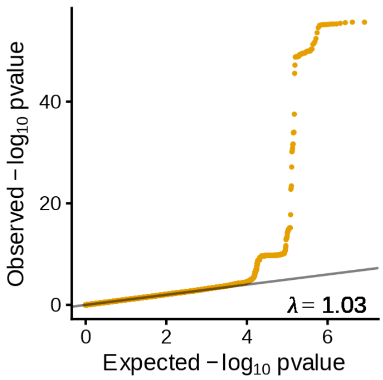


**Supplementary Figure 1. Quantile-quantile plot for the GWAS of neonatal jaundice.** The GWAS is based on the neonate's genome (n = 27,384 neonates, cases = 1,826), and observed p-values were two-sided.

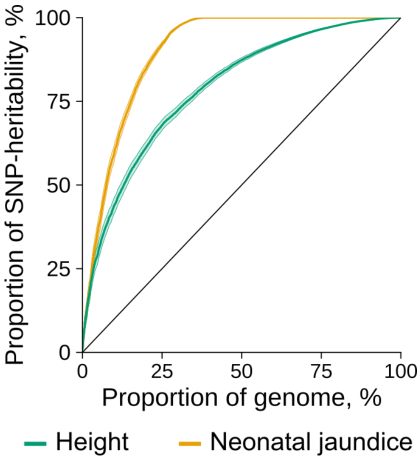


**Supplementary Figure 2. Contrast of polygenicity of neonatal jaundice and adult height.** We estimated local heritability using Heritability Estimation from Summary Statistics (HESS) for neonatal jaundice and adult height. We first sorted all regions by proportion of heritability and then calculated the cumulative proportion of the genome (x-axis), and the cumulative proportion of heritability (y-axis). The identity line indicates an imaginary polygenic trait with equal contributions from all variants, and the band surrounding the estimate, the 95% CI.

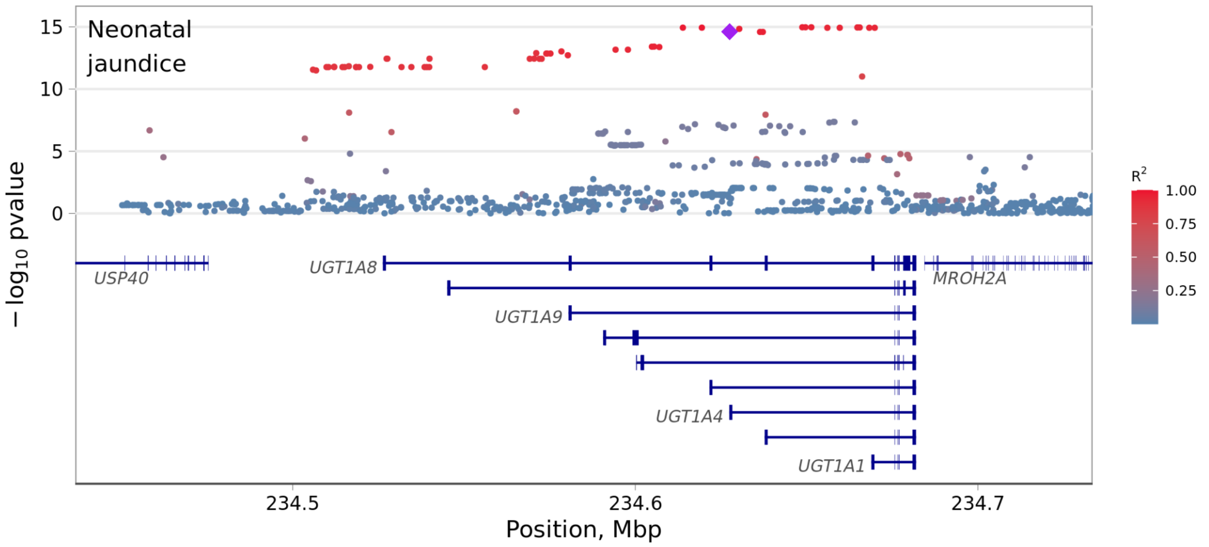


**Supplementary Figure 3. UGT1A locus in the replication cohort from Denmark.** Y-axis shows the two-sided p-value of the associations between the variants and neonatal jaundice (replication cohort, neonatal genome, n = 6,902, cases = 1,300). Highlighted are the missense variant (rs6755571, diamond) and variants in LD with it. LD was estimated in 23,196 non-related mothers from MoBa.

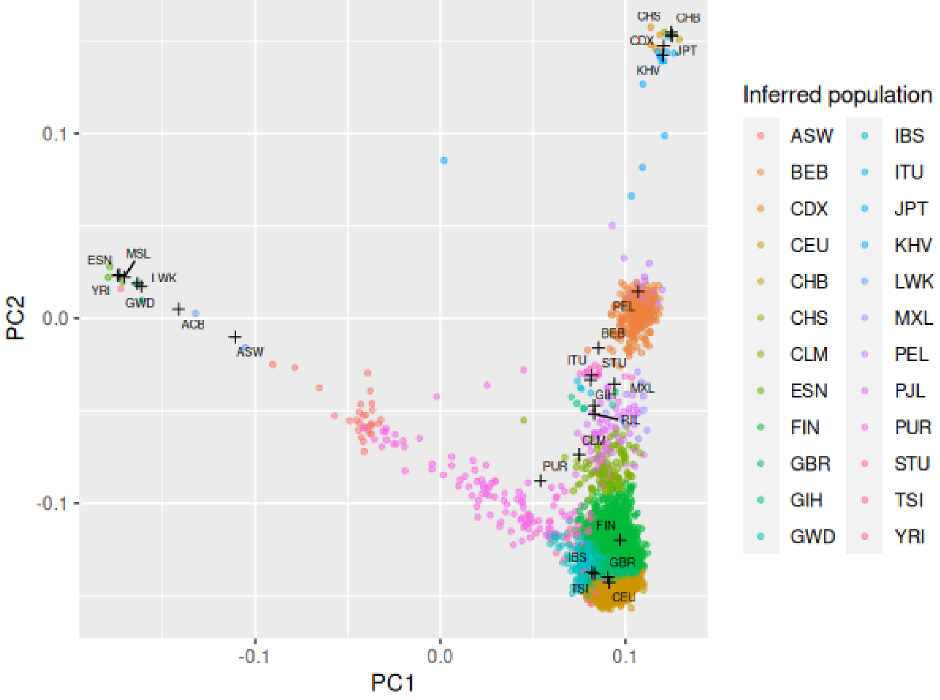


**Supplementary Figure 4. Inferred ancestries of the neonatal samples in MoBa.** MoBa sample ancestry was inferred with a nearest centroid classifier (in the space of the first 3 PCs), trained on populations from the 1000 Genomes Project data: crosses show the centroids of each population in 1000 Genomes, and neonatal samples (points, n = 28,112) are assigned to the nearest population by Euclidean distance to the centroid. Shown here is the projection of the points and centroids to PCs 1 and 2.

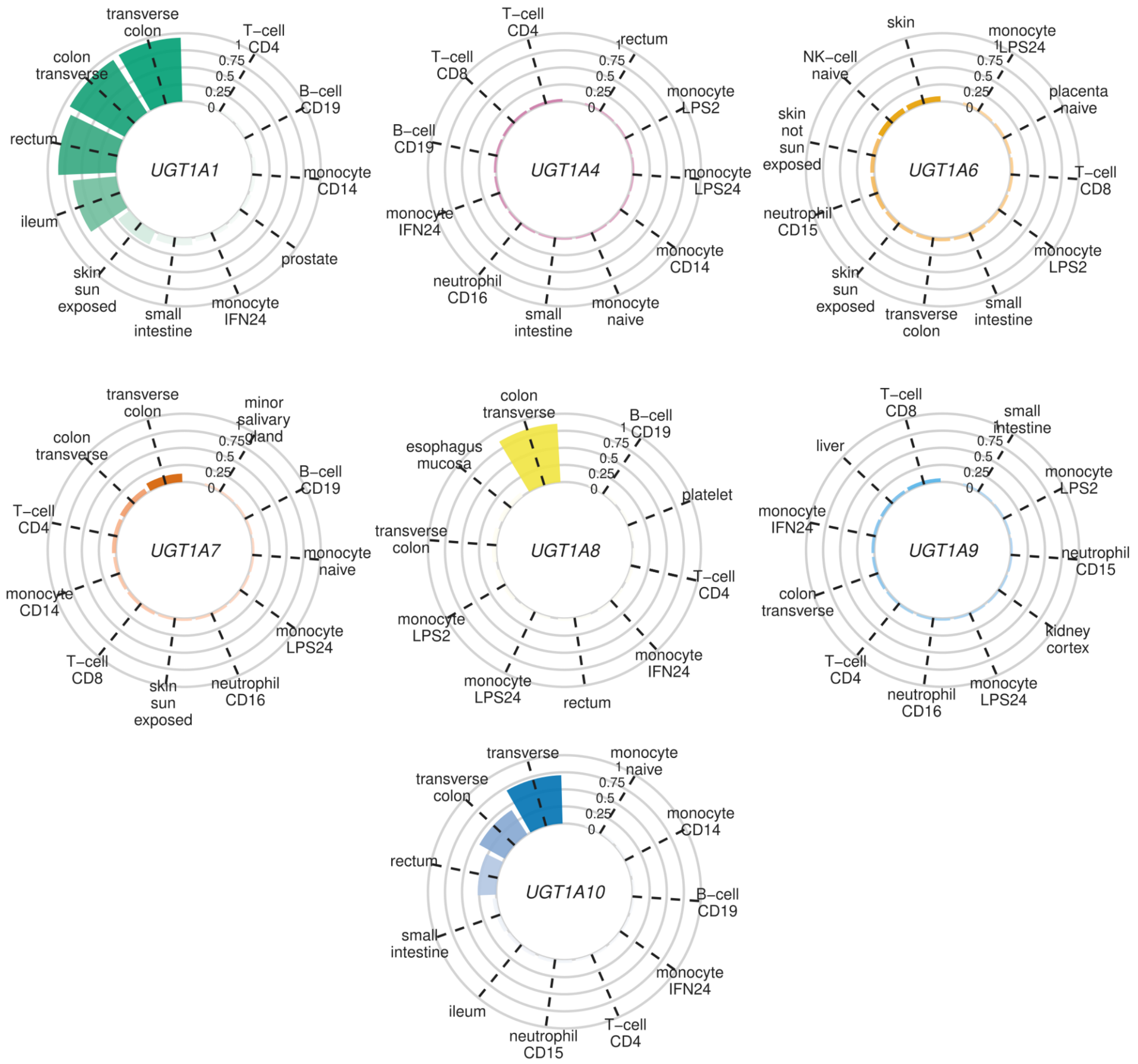


**Supplementary Figure 5. Colocalization between neonatal jaundice and cis-eQTLs for *UGT1A** genes.** We performed colocalization analyses between our GWAS of neonatal jaundice (n = 27,384 neonates, cases = 1,826) and cis-eQTLs from 177 different cell types/ tissues from the eQTL Catalogue for seven *UGT1A** genes. For each gene, we show only the top ten cell types/ tissues that had the largest probability of colocalization with neonatal jaundice, with values ranging from 0-1.

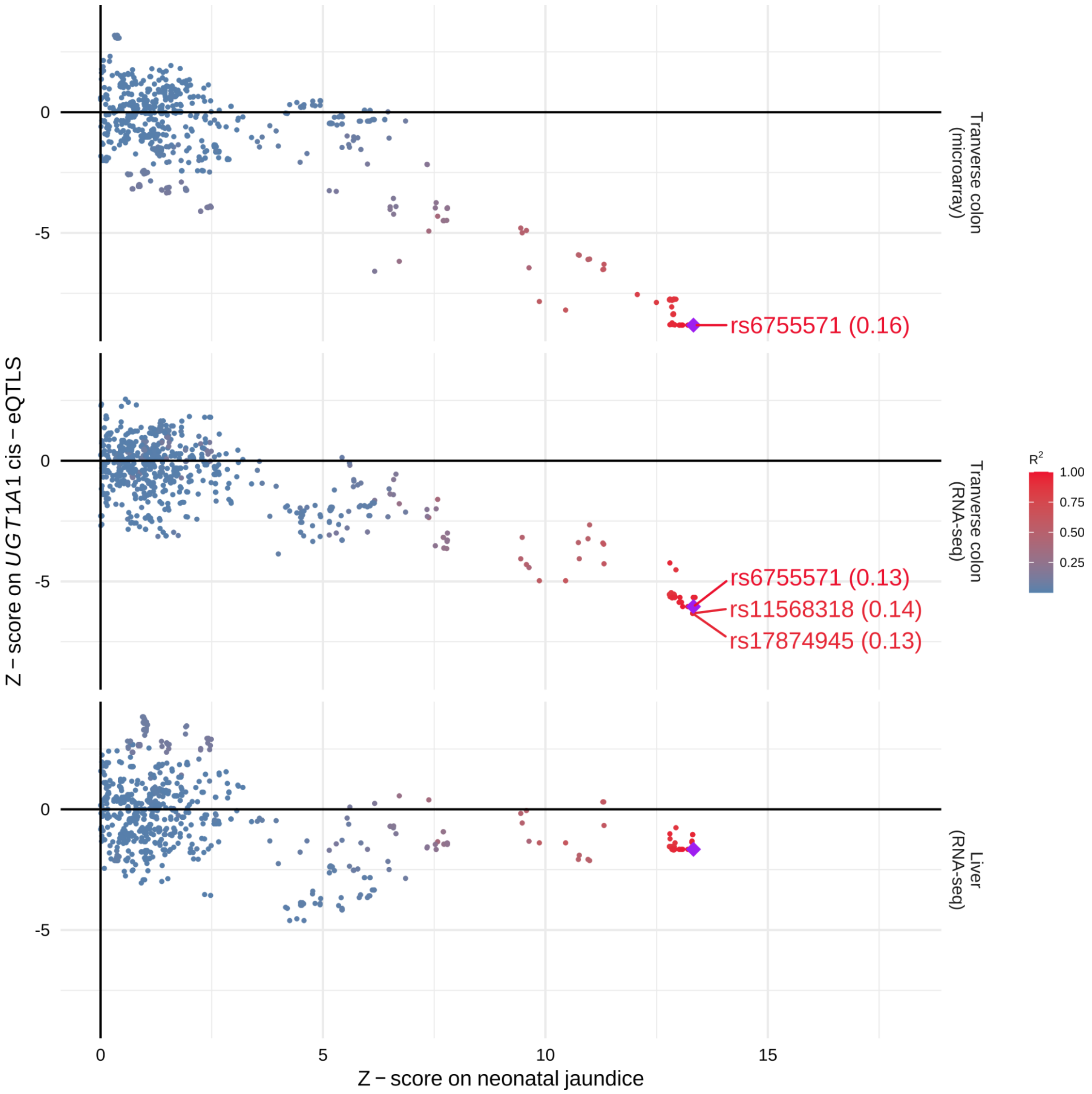


**Supplementary Figure 6. Effects on neonatal jaundice and cis-eQTLs of UGT1A1 in the colon and liver.** We performed colocalization analyses between our GWAS of neonatal jaundice (n = 27,384 neonates, cases = 1,826) and cis-eQTLs from 177 different cell types/ tissues from the eQTL Catalogue for seven *UGT1A** genes. Here, we show the correlation between the z-scores on neonatal jaundice and those on eQTLs of UGT1A1 in the colon (microarray and RNA-seq) and liver. A shared causal variant between neonatal jaundice and eQTLs of UGT1A1 in the colon best explained the associations in this locus (posterior probability of colocalization > 0.9); we show data from the liver even though we observed no colocalization. The SNPs with a posterior probability of colocalization > 0.1 are highlighted (only for eQTLs that colocalized with neonatal jaundice), as is the missense variant (rs6755571, diamond) and variants in LD with it. LD was estimated in 23,196 non-related mothers from MoBa.

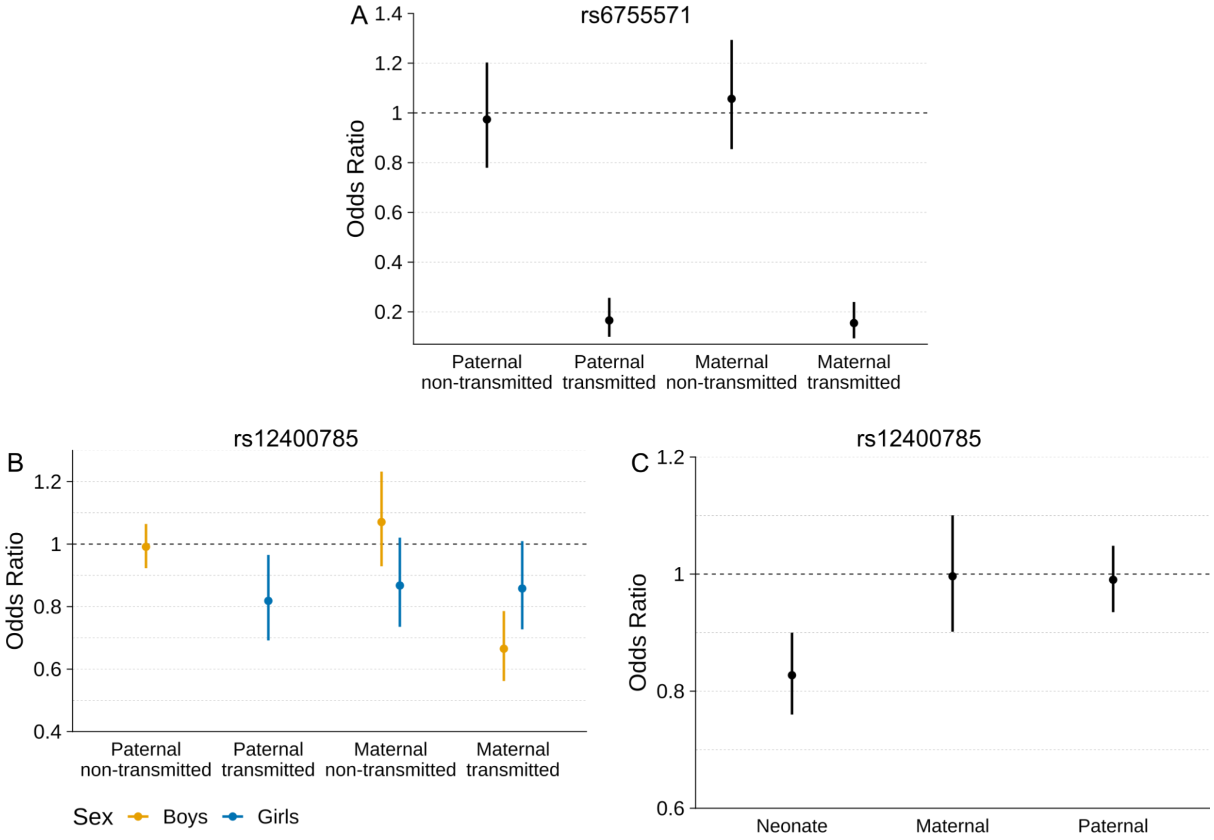


**Supplementary Figure 7. Analysis of the parental transmitted and non-transmitted alleles of rs6755571 and rs12400785 identified in the GWAS of neonatal jaundice (neonatal genome).** In all panels, the dot represents the odds ratio for the association, and the error bars the 95% CI estimated using logistic models. A, association between parental transmitted and non-transmitted alleles of rs6755571 (*UGT1A** genes region) and neonatal jaundice (n = 23,196 parent-offspring, n cases = 1,569). B, association between parental transmitted and non-transmitted alleles of rs12400785 and neonatal jaundice. This locus is located at the X chromosome, so the analysis was conducted in girls (maternal transmitted and non-transmitted and paternal transmitted, n = 11,438 parent-offspring, n cases = 686) and boys (maternal transmitted and non-transmitted and paternal non-transmitted, n = 11,758 parent-offspring, n cases = 883) separately. C, conditional analysis between neonate, maternal and paternal rs12400785 dosages and neonatal jaundice (n = 23,196 parent-offspring, n cases = 1,569). This locus is located on the X chromosome (males were coded as diploid).

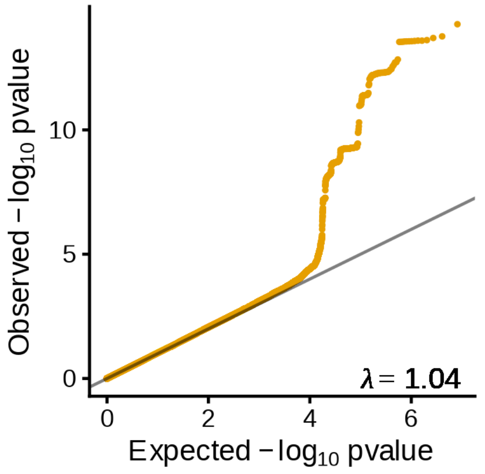


**Supplementary Figure 8. Quantile-quantile plot for the GWAS of offspring neonatal jaundice.** The GWAS is based on the maternal genome (n = 29,182, n cases = 2,401), and observed p-values were two-sided.

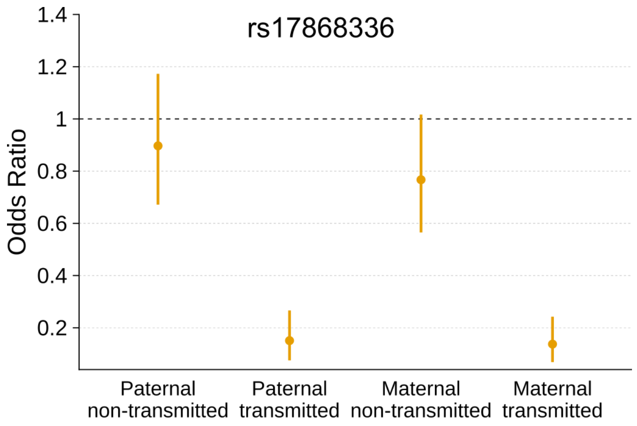


**Supplementary Figure 9. Effect of the parental transmitted and non-transmitted alleles of rs17868336 identified in the GWAS of offspring neonatal jaundice (maternal genome).** Association between parental transmitted and non-transmitted alleles of rs6755571 (*UGT1A** genes region) and neonatal jaundice (n = 23,196 parent-offspring trios, cases = 1,569). The dot represents the odds ratio for the association, and the error bars the 95% CI.

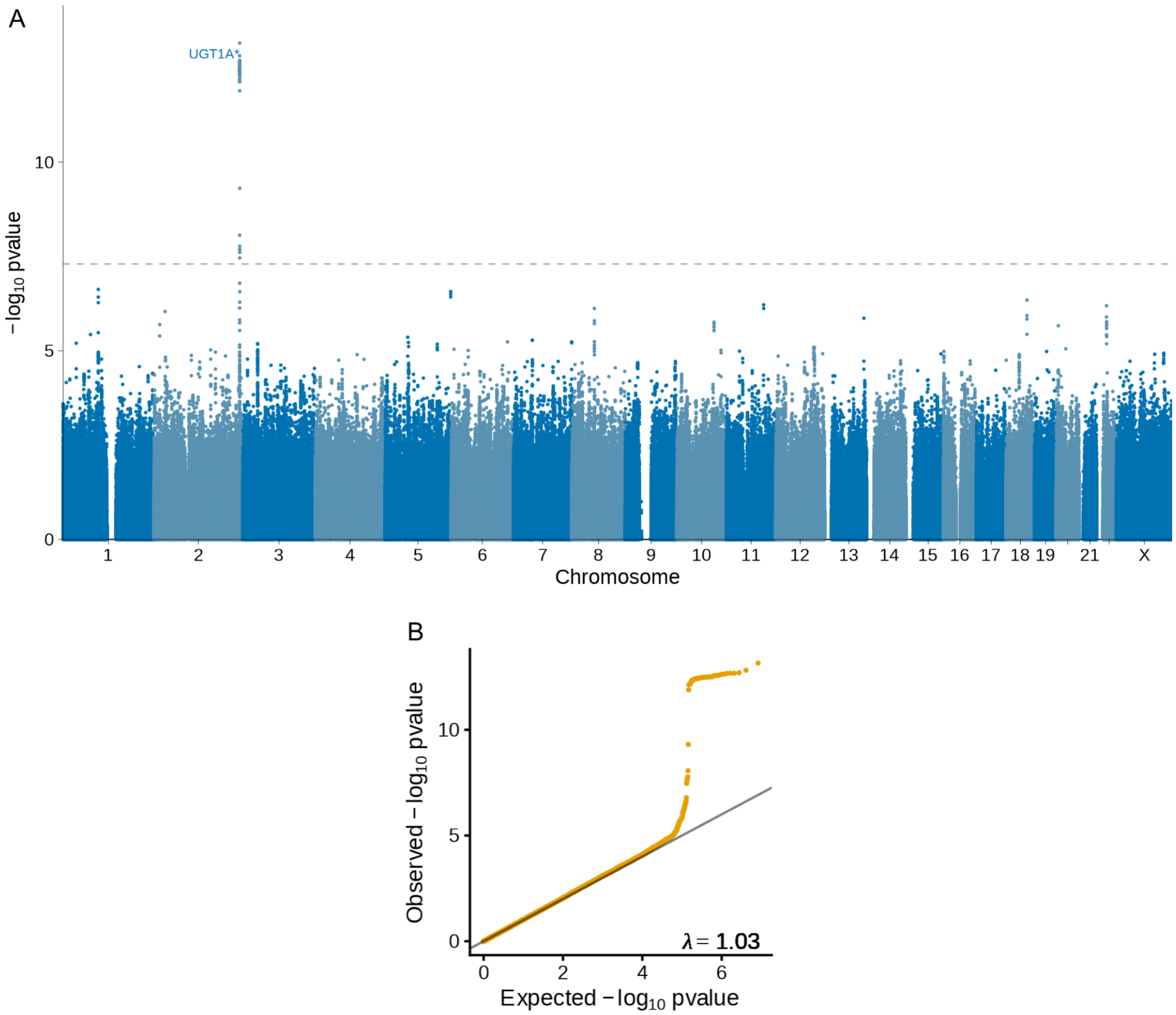


**Supplementary Figure 10. Manhattan and quantile-quantile plot of the GWAS of offspring neonatal jaundice using the paternal genome.** (A) Manhattan plot of and neonatal jaundice (n = 28,384, n cases = 2,361). The x-axis shows the chromosome position and the y-axis the two-sided p-value of the association. The dashed line represents the genome-wide significance threshold (p-value = $5 \times 10^{-8}$). Each genome-wide significant locus is labeled by their nearest protein-coding gene. (B) Quantile-quantile plot of the GWAS of offspring neonatal jaundice using the paternal genome (n = 28,384, n cases = 2,361, observed p-values are two-sided).

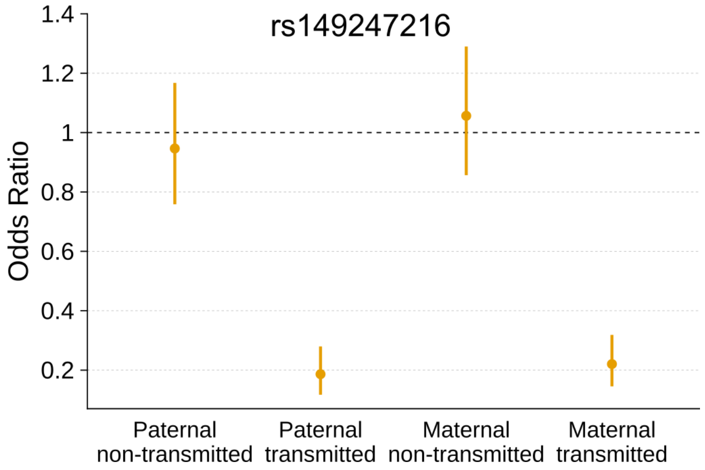


**Supplementary Figure 11. Effect of the parental transmitted and non-transmitted alleles of rs149247216 identified in the GWAS of offspring neonatal jaundice (paternal genome).** Association between parental transmitted and non-transmitted alleles of rs149247216 (*UGT1A** genes region) and neonatal jaundice (n = 23,196 parent-offspring trios, cases = 1,569). The dot represents the odds ratio for the association, and the error bars the 95% CI.

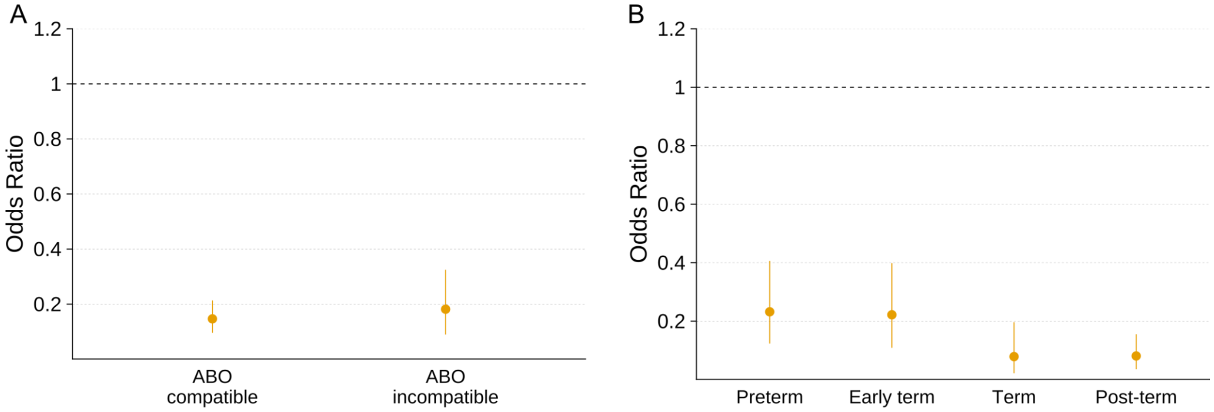


**Supplementary Figure 12. Interactions between the neonatal missense variant (rs6755571) and maternal-fetal blood group incompatibility and gestational duration.** In all panels, the dot represents the odds ratio for the association on neonatal jaundice, and the error bars the 95% CI. (A) Interaction between neonatal rs6755571 and maternal-fetal ABO blood group incompatibility (n = 23,196 parent-offspring trios, cases = 1,569, p-value = 0.809). ABO blood groups in mothers and neonates were determined genetically, and ABO incompatibility was defined as maternal O blood group and neonate A or B. (B) Interaction between neonatal rs6755571 and gestational duration (n = 23,196 parent-offspring trios, cases = 1,569, p-value = $6.2 \times 10^{-4}$). For visualization purposes, we have split gestational duration into preterm (below 259 days, n = 737), early term (between 259 and 273 days, n = 3389), term (between 273 and 280 days, n = 5,161) and post-term (above 280 days, n = 13,675) deliveries.

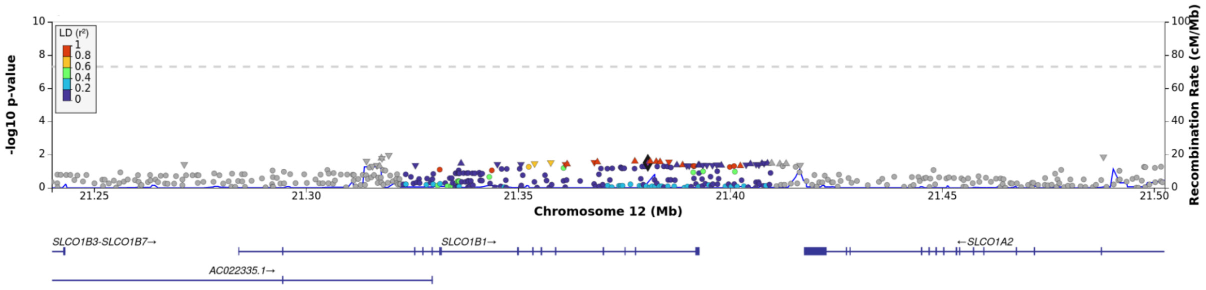



**Supplementary Figure 13.** Locus Zoom plot of the GWAS of neonatal jaundice at *SLCO1B1* gene region. This region has been previously found to be associated with adult bilirubin levels, but is not with neonatal jaundice (n = 27,384 neonates, cases = 1,826). The lead variant of adult bilirubin levels in this locus (rs4149081, diamond) and the variants in LD with it are highlighted. The x-axis shows the chromosome position and the y-axis the two-sided p-value.

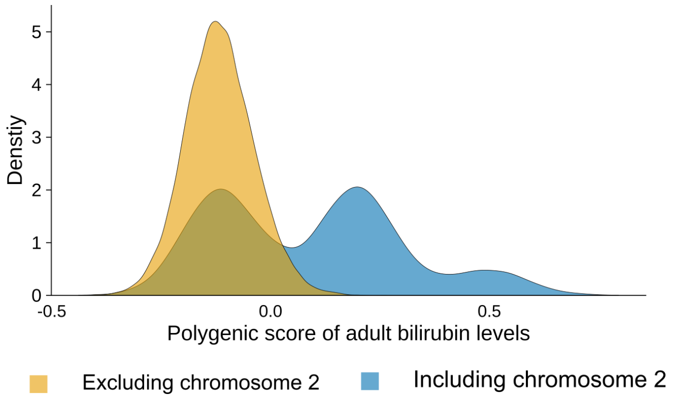



**Supplementary Figure 14. Distribution of the polygenic score of adult bilirubin levels with or without including chromosome 2.** The polygenic score was constructed using weights previously deposited at the PGS Catalog (ID: PGS002160). n = 23,196 neonates
